# Prevalence of dysphagia in patients with multiple sclerosis; a systematic review and meta-analysis

**DOI:** 10.1101/2021.06.07.21258473

**Authors:** Omid Mirmosayyeb, Narges Ebrahimi, Arman Shekarian, Sara Bagherieh, Alireza Afshari-Safavi, Vahid Shaygannejad, Mahdi Barzegar

**Affiliations:** Department of Neurology, School of Medicine, Isfahan University of Medical Sciences, Isfahan, Iran; Isfahan Neurosciences Research Center, Isfahan University of Medical Sciences, Isfahan, Iran; Department of Biostatistics and Epidemiology, Faculty of Health, North Khorasan University of Medical Sciences, Bojnurd, Iran

**Keywords:** Multiple Sclerosis, Demyelinating Diseases, Autoimmune Diseases, Dysphagia, Prevalence

## Abstract

**Objectives:** Dysphagia is a major comorbidity observed in patients with multiple sclerosis, yet different prevalence rates are reported for it. Therefore, we have designed this systematic review to estimate the pooled prevalence of dysphagia in patients with MS.

**Method:** We searched PubMed, Scopus, EMBASE, Web of Science, and gray literature including references from the identified studies, reviews studies, and conference abstracts which were published up to February 2021. Articles that were relevant to our topic and could provide information regarding the prevalence of dysphagia among MS patients were included; however, articles with self-report screening strategies were excluded.

**Results:** The literature search found 1993 articles. After eliminating duplicates, 1272 articles remained. Sixteen abstract conference papers were included for final analysis. A total of 10846 MS cases and 4913 MS patients with dysphagia included in the analysis. The pooled prevalence of dysphagia in the included studies was 45.3% (95% CI: [40.7%-50%])

**Conclusion:** The results of this systematic review shows that the prevalence of dysphagia in MS patients is 45% which is greatly higher compared to the general population.

## Introduction

Multiple Sclerosis (MS) is an autoimmune disease caused by the demyelination of neurons within the Central Nervous System (CNS). It is generally more prevalent among women and younger people and deteriorates patients’ lifestyles in every manner ranging from personal, social, occupational, to marital aspects.(1,2) MS can be classified based on its clinical manifestation and course of progression, as there are many differences in the manifestation of MS between different patients of different ages, genders, races, genetic, and even geographical backgrounds. However, symptoms associated with MS ordinarily pivot around sensory and motor dysfunctions. Such dysfunctions include but are not limited sensory deficits such as vision, hearing, and olfaction loss, besides motor deficits being gastrointestinal, bladder-control, gait, and swallowing disorders.(3–5)

Swallowing or deglutition is a half-automated motor action that requires the involvement of respiratory, oropharyngeal, and gastrointestinal muscles. This process is thought to occur in four stages: oral preparatory, oral, pharyngeal, and esophageal stages. Any difficulty swallowing can be loosely termed “Dysphagia”. Dysphagia has been known as a clinical finding in MS since as early as the 19th century.(6,7) The prevalence of dysphagia among MS patients has in fact been shown to be higher than that of the normal population. Swallowing is an orchestration of numerous muscles, cranial nerves, and neural pathways and MS lesions can affect the process in any of the four stages, leading to disturbances in this mechanism that can cause clinical symptoms such as weight loss, dehydration, halitosis, and even aspiration pneumonia in the affected MS patients.(8) Such patients are increasingly prone to developing severe dysphagia as it is a frequently overlooked symptom among physicians and is almost always under-reported unless it co-occurs with episodes of aspiration pneumonia that is caused when a damaged control mechanism, due to dysphagia, allows food to be misled to the respiratory tract. Identifying dysphagia in the early stages can help reduce its consequences and apply better preventive measures to stop the progression of symptoms in those at risk.(4,9)

Regarding the growing evidence on the higher prevalence of dysphagia among MS patients and the importance of dysphagia as a part of MS disease follow-up, the present study has been conducted to estimate the prevalence of dysphagia in MS patients and to assess the potential risk factors.

## Methods

### Literature search

We searched PubMed, Scopus, EMBASE, Web of Science, and gray literature including references from the identified studies, review studies, and conference abstracts which were published up to February 2021.

### Inclusion criteria

Studies reporting the prevalence of dysphagia among MS participants with a sample size of over at least 10 patients, for whom the diagnosis of dysphagia in MS patients was made using any strategy other than self-report methods were included. Nevertheless, case reports and case series articles, articles that were written in any language other than English, and any study that had used self-report screening methods as the means to diagnosing dysphagia among MS patients were excluded. Studies with unclear diagnostic methods were excluded as well.

### Data search and extraction

We conducted a systematic computerized search using four data banks: PubMed (NCBI), Scopus, web of science, and Embase. We also searched the gray literature including references from the identified studies, reviews studies, and conference abstracts which were published up to February 2021.

We used Mesh terms and text words to generate a syntax that included two components. First, we used “Deglutition Disorders”, “Deglutition Disorder”, (Disorders AND Deglutition), “Swallowing Disorders”, “Swallowing Disorder”, “Dysphagia”, “Oropharyngeal Dysphagia”, (Dysphagia AND Oropharyngeal), “Esophageal Dysphagia”, (Dysphagia AND Esophageal), (difficult* AND swallowing), and (difficult* AND deglutition) to identify the first component of our search and “Multiple Sclerosis”, (Sclerosis AND Multiple), (Sclerosis AND Disseminated), “Disseminated Sclerosis”, “MS, (Multiple Sclerosis)”, (“Multiple Sclerosis” AND “Acute Fulminating” were the words we used to identify the other search component.

Additionally, we customized our search syntax (query) for each data bank. Two researchers independently screened the articles. The following data was extracted from the included studies: first author, region, publication date, type of study, sample size of case and control, and the demographic variables for case and control such as sex and mean age. Other variables that we collected in our table included the exact name of the dysphagia diagnostic test which included instrumental techniques, clinical examinations, screening strategies or other techniques, MS subtype, disease duration, EDSS score, number of slight, alarming, moderate, and severe dysphagia in both case and control. Besides, number of dysphagia was reported in categories as follows: different MS subtypes, solid and liquid dysphagia, oropharyngeal, oral, pharyngeal, esophageal, neurogenic, and functional dysphagia. Moreover, number of aspirations due to dysphagia was among the variables that we extracted from articles.

### Statistical analysis

Meta-analysis was performed using STATA 14 software (STATA Corporation, College Station, Texas, USA). A forest plot was applied to show the prevalence of dysphagia across all included studies and the pooled prevalence with corresponding 95% confidence intervals (95% CI). The Cochran’s chi-square test and inconsistency index (I^2^) were used to check between study heterogeneity. If the I^2^ statistics was greater than 75% (high heterogeneity), a random effect model was done using DerSimonian and Larid approach. Subgroup analysis was conducted by diagnostic test method (instrumental strategies, screening strategies, clinical examination strategies and other strategies), sample size (≤ 100 and > 100), publication year (≤ 2010 and > 2010), EDSS (≤ 4 and > 4) and disease duration (≤ 10 and > 10). A funnel plot of logit transformed prevalence was applied to investigate publication bias with Egger regression asymmetry and Begg’s tests. If evidence of publication bias was observed, then trim and fill technique was used to adjust the effects. Level of statistical significance for all tests was considered to be less than 0.05.

## Results

The literature search found 1993 articles. After eliminating duplicates, 1272 articles remained. Sixteen abstract conference papers were included for final analysis. A total of 10846 MS cases and 4913 MS patients with dysphagia included in the analysis. The pooled prevalence of dysphagia in the included studies was 45.3% (95% CI: [40.7%-50%]) (Figure 1)

**Figure 1.**
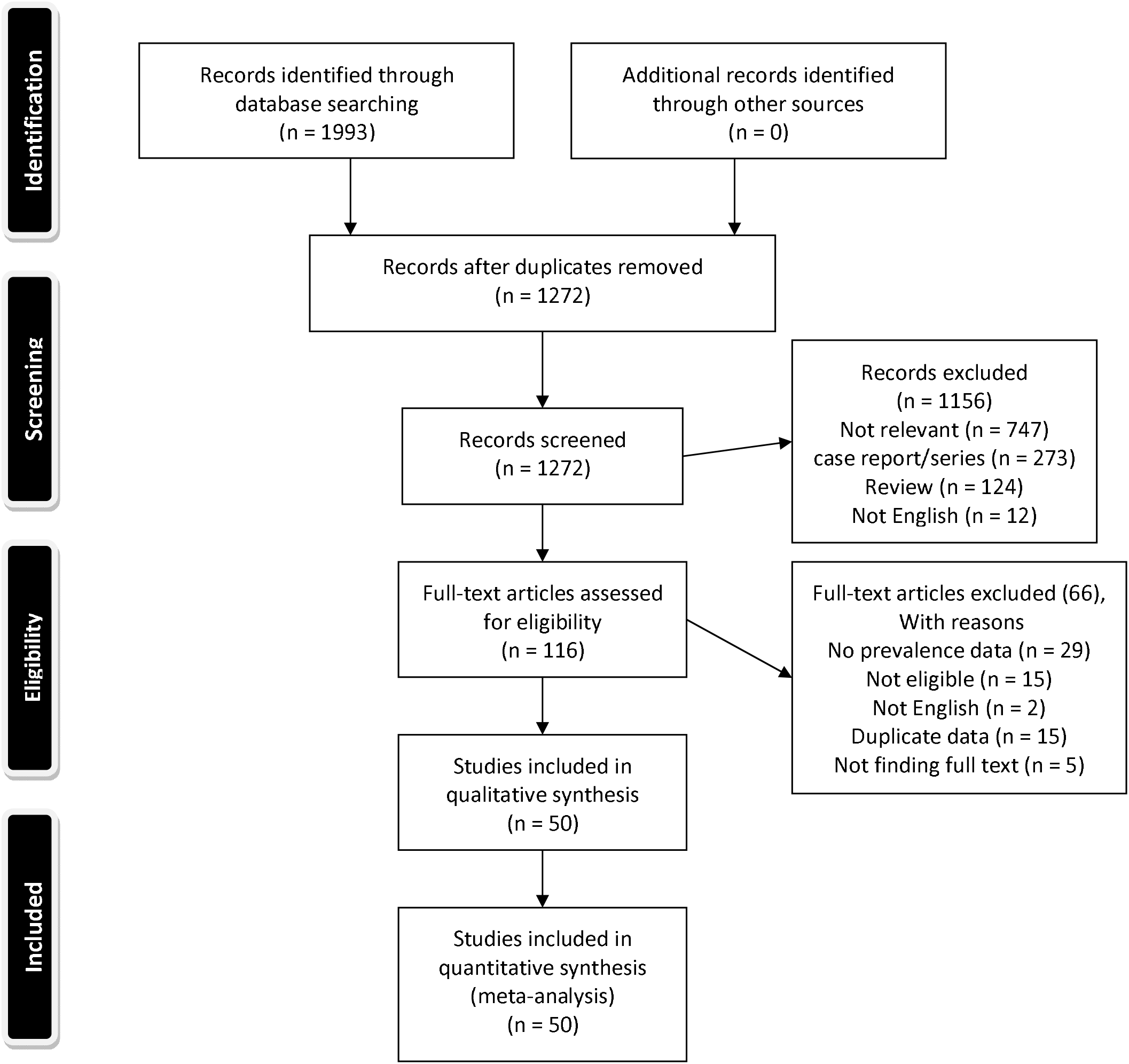
Flow diagram summarizing the selection of eligible studies.

### Critical appraisal

The quality of all the included articles was assessed using the Joanna Briggs Institute (JBI) critical appraisal checklist. The JBI checklist is the preferred tool for measuring the quality of descriptive studies reporting prevalence data and has a system of ranking articles based on the number of “YES” answers they earn according to its questions. The number of “YES” answers an article can earn ranges between 0 to 9.(10) Using this checklist, 8 Of the included studies earned less than 4 “YES” answers, 22 studies earned between 4 to 6 “YES” answers, and 4 studies earned more than 6 “YES” answers. (Supplementary *1*)

**Supplementary 1.** Table of quality assessment using the JBI checklist

|  | Q1<br>Was the sample frame appropriate to address the target population? | Q2<br>Were study participants sampled in an appropriate way? | Q3<br>Was the sample size adequate? | Q4<br>Were the study subjects and the setting described in detail? | Q5<br>Was the data analysis conducted with sufficient coverage of the identified sample? | Q6<br>Were valid methods used for the identification of the condition? | Q7<br>Was the condition measured in a standard, reliable way for all participants? | Q8<br>Was there appropriate statistical analysis? | Q9<br>Was the response rate adequate, and if not, was the low response rate managed appropriately? |
| --- | --- | --- | --- | --- | --- | --- | --- | --- | --- |
| Levinthal,2013 | NO | NO | YES | NO | UNCLEAR | YES | YES | YES | UNCLEAR |
| Danesh Pajouh, 2017 | NO | NO | YES | YES | UNCLEAR | YES | YES | YES | UNCLEAR |
| Thomas,1999 | NO | NO | NO | YES | UNCLEAR | YES | YES | YES | UNCLEAR |
| Calcagno, 2002 | NO | NO | YES | YES | UNCLEAR | YES | YES | YES | UNCLEAR |
| DePauw, 2002 | YES | NO | YES | YES | UNCLEAR | YES | YES | YES | UNCLEAR |
| Printza, 2019 | NO | NO | YES | YES | UNCLEAR | YES | YES | YES | UNCLEAR |
| Santos, 2019 | NO | NO | NO | YES | UNCLEAR | YES | YES | YES | UNCLEAR |
| Beckmann, 2015 | UNCLEAR | UNCLEAR | NO | NO | UNCLEAR | YES | YES | YES | UNCLEAR |
| Alfonsi, 2013 | NO | NO | NO | NO | UNCLEAR | YES | YES | YES | UNCLEAR |
| Di Stadio, 2021 | NO | NO | NO | YES | YES | YES | YES | YES | YES |
| Covello, 2020 | NO | UNCLEAR | YES | NO | UNCLEAR | YES | YES | YES | UNCLEAR |
| Abraham, 1997 | NO | NO | YES | NO | YES | UNCLEAR | YES | YES | UNCLEAR |
| Goncalves, 2016 | NO | NO | NO | NO | UNCLEAR | YES | YES | YES | UNCLEAR |
| Poorjavad, 2010 | NO | NO | YES | YES | UNCLEAR | YES | YES | YES | UNCLEAR |
| Fernandes, 2013 | NO | UNCLEAR | YES | NO | UNCLEAR | YES | YES | YES | UNCLEAR |
| Klugman, 2002 | YES | NO | NO | NO | NO | UNCLEAR | YES | YES | UNCLEAR |
| Tarameshlu, 2017 | YES | NO | YES | YES | NO | YES | YES | YES | UNCLEAR |
| Solaro, 2019 | YES | NO | YES | YES | YES | YES | YES | YES | UNCLEAR |
| Solaro, 2013 | YES | YES | YES | YES | YES | YES | YES | YES | UNCLEAR |
| Tahir, 2020 | NO | NO | NO | NO | NO | YES | YES | YES | UNCLEAR |
| Johansson, 2020 | YES | NO | YES | NO | NO | UNCLEAR | YES | YES | UNCLEAR |
| Hartelius, 1994 | NO | UNCLEAR | YES | NO | NO | NO | YES | YES | UNCLEAR |
| Wiesner, 2001 | NO | NO | UNCLEAR | NO | NO | YES | YES | YES | UNCLEAR |
| Sadeghi, 2020 | NO | NO | YES | YES | UNCLEAR | YES | YES | YES | UNCLEAR |
| Bergamaschi, 2008 | YES | NO | YES | YES | YES | YES | YES | YES | UNCLEAR |
| Alali, 2018 | YES | YES | YES | YES | UNCLEAR | YES | YES | YES | UNCLEAR |
| Printza, 2020 | NO | NO | YES | YES | UNCLEAR | YES | YES | YES | UNCLEAR |
| Milewska, 2020 | NO | NO | NO | NO | UNCLEAR | YES | YES | YES | UNCLEAR |
| Sales, 2013 | NO | NO | YES | YES | UNCLEAR | YES | YES | YES | UNCLEAR |
| Bergamaschi, 2009 | YES | NO | YES | YES | UNCLEAR | YES | YES | YES | UNCLEAR |
| Jafari, 2018 | NO | NO | YES | YES | UNCLEAR | YES | YES | YES | UNCLEAR |
| Tenekeci, 2018 | NO | NO | YES | NO | UNCLEAR | YES | YES | YES | UNCLEAR |
| Barzegar, 2021 | NO | UNCLEAR | YES | NO | UNCLEAR | YES | YES | YES | UNCLEAR |
| Danesh-Sani, 2013 | YES | NO | YES | NO | UNCLEAR | YES | YES | YES | UNCLEAR |

### Prevalence estimates

The overall prevalence of dysphagia across all 57 studies was 45.3% (95% CI: [40.7%-50%]), with a high level of heterogeneity (Q=; I^2^=95.17%; p<0.001). The highest and lowest prevalence of dysphagia was reported by Zenginler et al. (prevalence=100%; 95% CI: [88.8%-100%]) and Almerie et al. (prevalence=9%; 95% CI: [4%-16.9%]), respectively.

### Subgroup analysis

The prevalence estimates are presented by dysphagia diagnostic test (instrumental strategies, screening strategies, clinical examination strategies and other strategies) in Figure 2, Figure 3, Figure 4, and Figure 5. There was high heterogeneity in all the four analyses (I^2^= 94.89%, 94.15%, 93.65% and 88.73%, respectively). The pooled prevalence of dysphagia based on instrumental strategies (prevalence=65.7%; 95% CI: [51.3%-78.9%]) was higher than screening strategies (prevalence=44.1%; 95% CI: [38.9%-49.3%]), clinical examination strategies (prevalence=38.4%; 95% CI: [21.6%-56.6%]) and other strategies (prevalence=29%; 95% CI: [22.1%-36.4%]).

**Figure 2.**
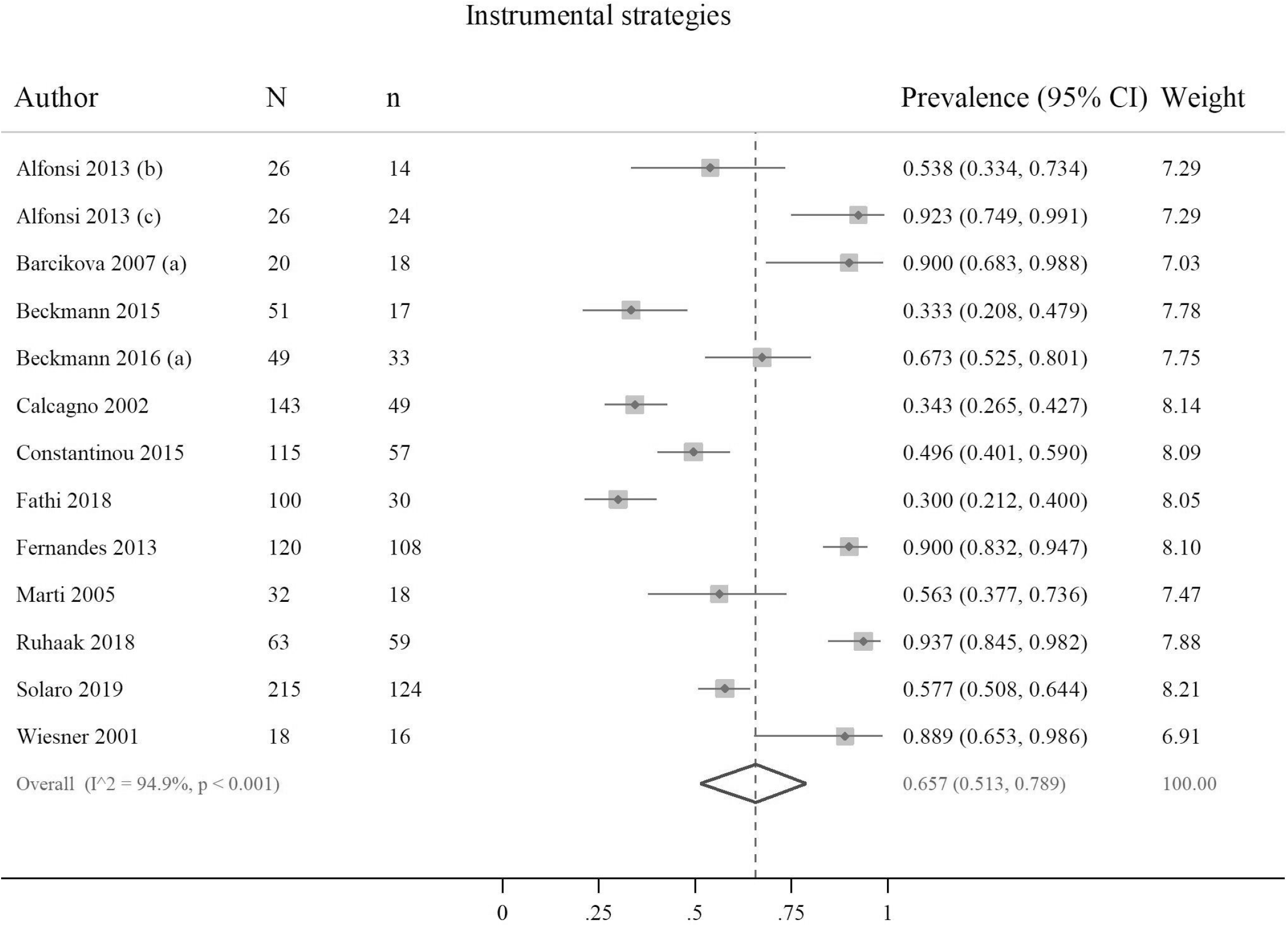
Prevalence of dysphagia based on instrumental strategies.

**Figure 3.**
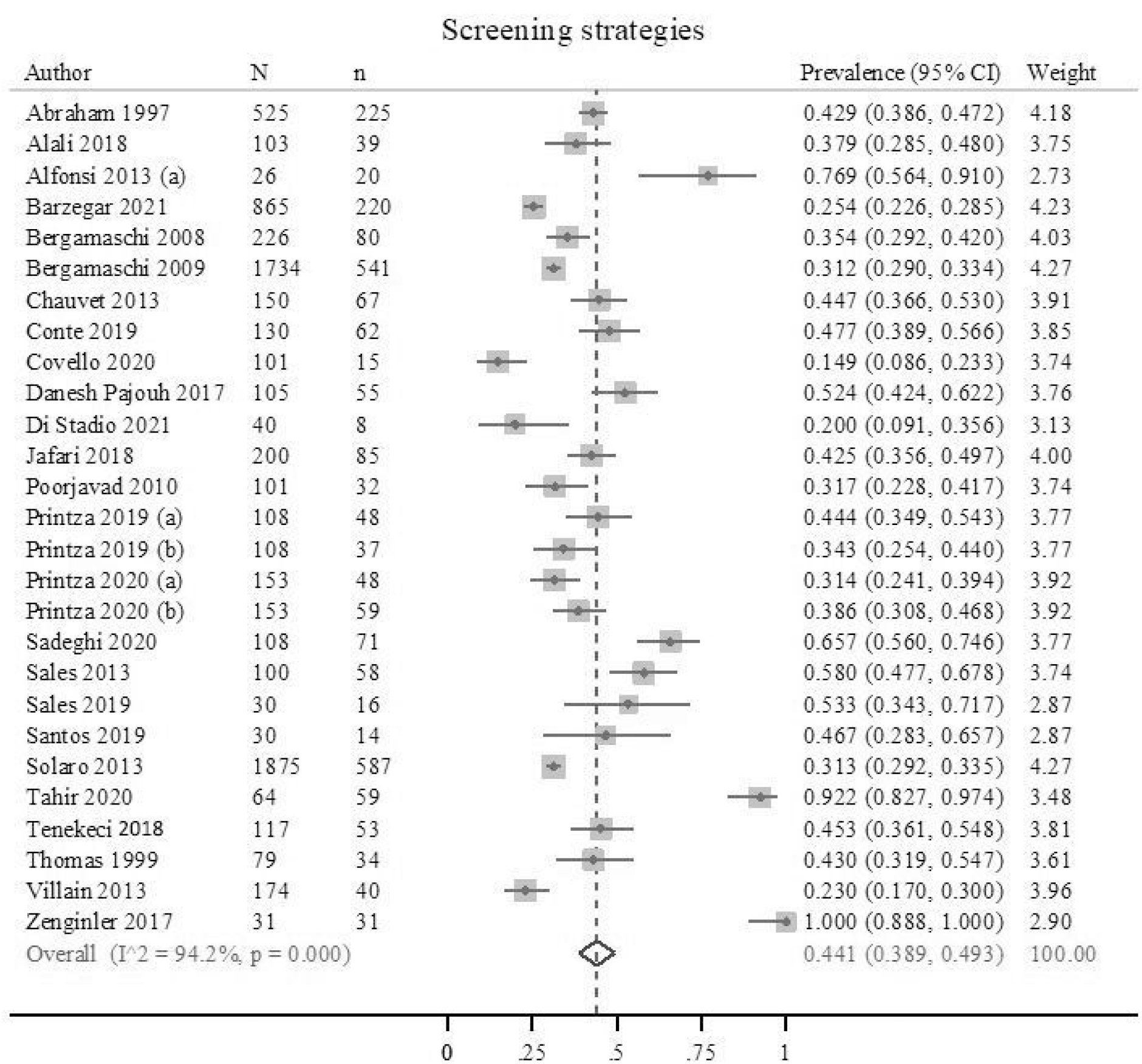
Prevalence of dysphagia based on screening strategies.

**Figure 4.**
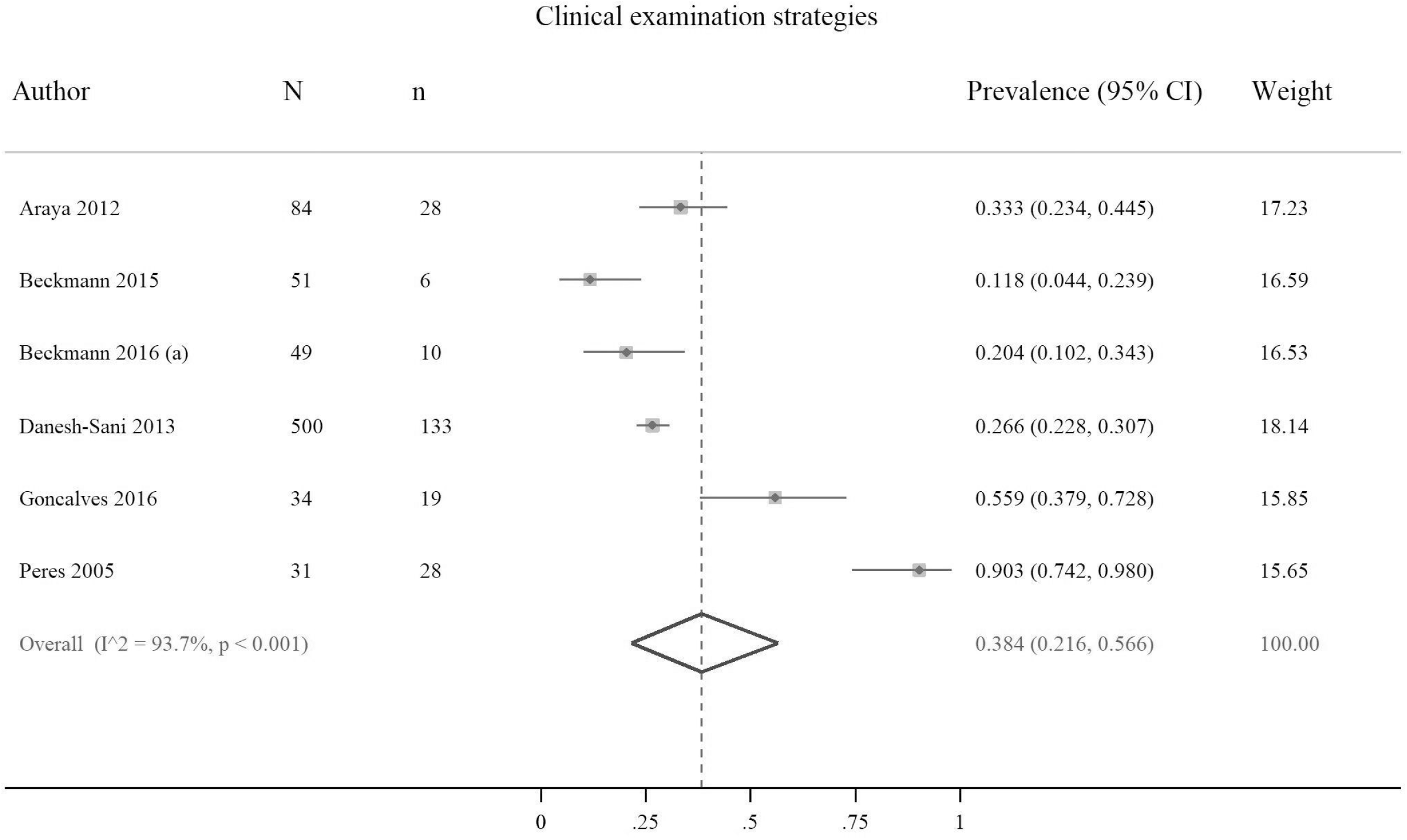
Prevalence of dysphagia based on clinical examination strategies.

**Figure 5.**
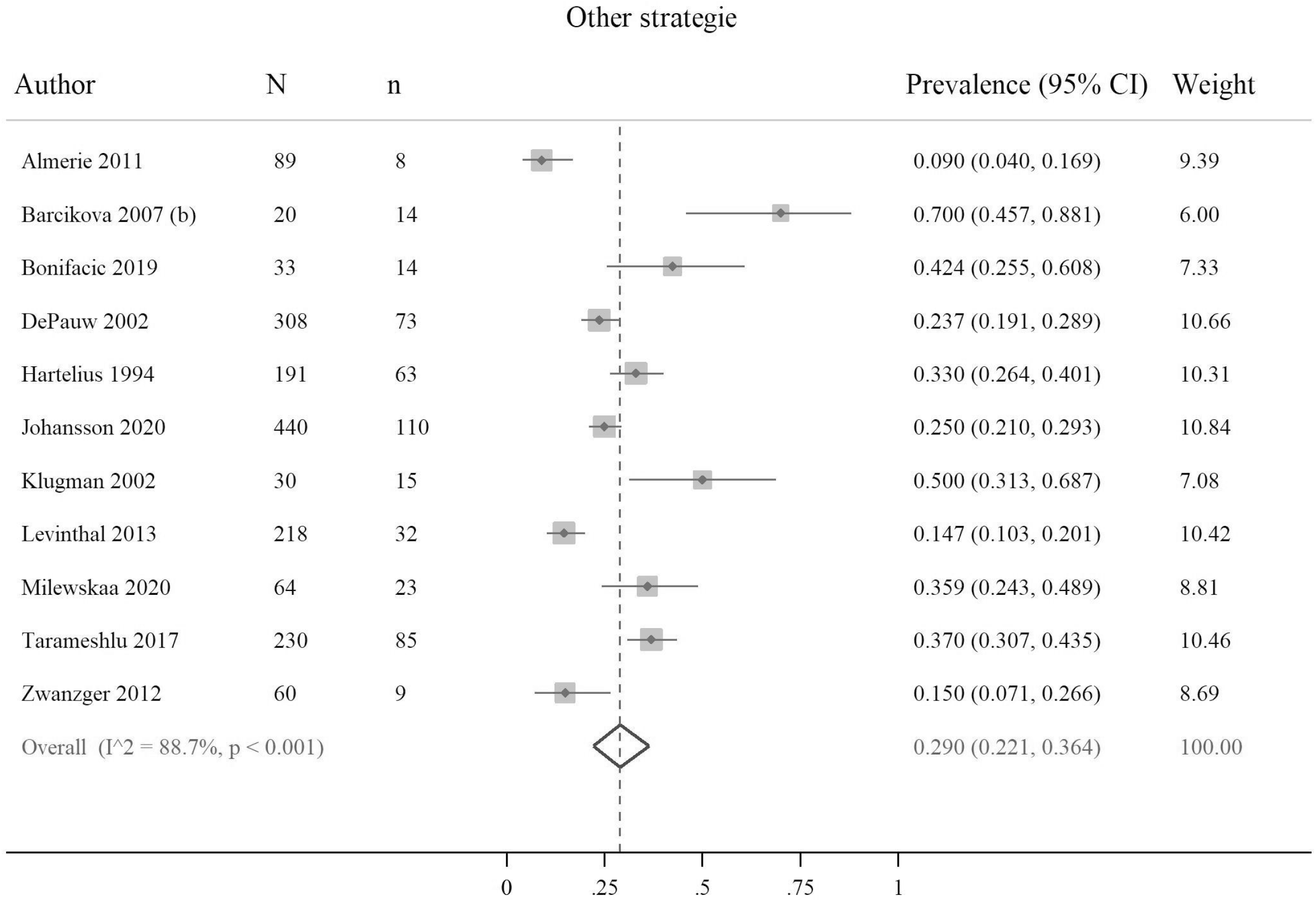
Prevalence of dysphagia based on other strategies, including non-instrumental strategies and any other method except for the instrumental, clinical, and screening strategies.

Subgroup analysis using, sample size, publication year, EDSS and disease duration was reported in Table 1. The pooled prevalence of dysphagia was higher in studies with a sample size less than 100 compared to those sample size greater than 100 (55.5% vs. 37.8%, p=0.019). However, prevalence of dysphagia was not significantly different in terms of publication year (47.5% vs. 44.3%, p=0.836), EDSS (46.8% vs. 58.2%, p=0.255) and disease duration (41% vs. 37.3%, p=0.585).

**Table 1.** Subgroup analysis of pooled prevalence of dysphagia

| **Table 1: Subgroup analysis of pooled prevalence of dysphagia** |  |  |  |  |  |  |
| --- | --- | --- | --- | --- | --- | --- |
| Subgroup by | | No. of | Total | Pooled prevalence | Heterogeneity $I^2$ | p-value |

Table 1 Subgroup analysis of pooled prevalence of dysphagia
|  |  | studies | sample size | (95% CI) |  |  |
| --- | --- | --- | --- | --- | --- | --- |
| Sample size | ≤ 100 | 28 | 1330 | 55.5% (43.4%-67.3%) | 94.76 | 0.019 |
|  | > 100 | 29 | 9516 | 37.8% (33.4%-42.3%) | 94.67 |  |
| publication year | ≤ 2010 | 14 | 3458 | 47.5% (40.1%-54.9%) | 92.03 | 0.836 |
|  | > 2010 | 43 | 7388 | 44.3% (38.4%-50.3%) | 95.78 |  |
| EDSS | ≤ 4 | 19 | 5547 | 46.8% (39.5%-54.1%) | 95.74 | 0.255 |
|  | > 4 | 13 | 1674 | 58.2% (48%-68.1%) | 92.88 |  |
| Disease duration | ≤ 10 | 10 | 1800 | 41% (29.7%-52.7%) | 94.98 | 0.585 |
|  | > 10 | 16 | 6034 | 37.3% (31.4%-43.4%) | 94.78 |  |

### Publication bias

No publication bias was found among the studies that were based on the instrumental strategies (bias= 4.01; p=0.051), clinical examination strategies (bias= 2.13; p=0.394) and other strategies (bias= 0.884; p=0.669), as depicted by the funnel plot (Figure 5) and the results of Egger’s and Begg’s tests. However, the evidence of publication bias was observed among the screening strategies-based studies (bias= 3.02; p=0.005), but the trim and fill method showed no need for additional studies. (Table 2)

**Table 2.**
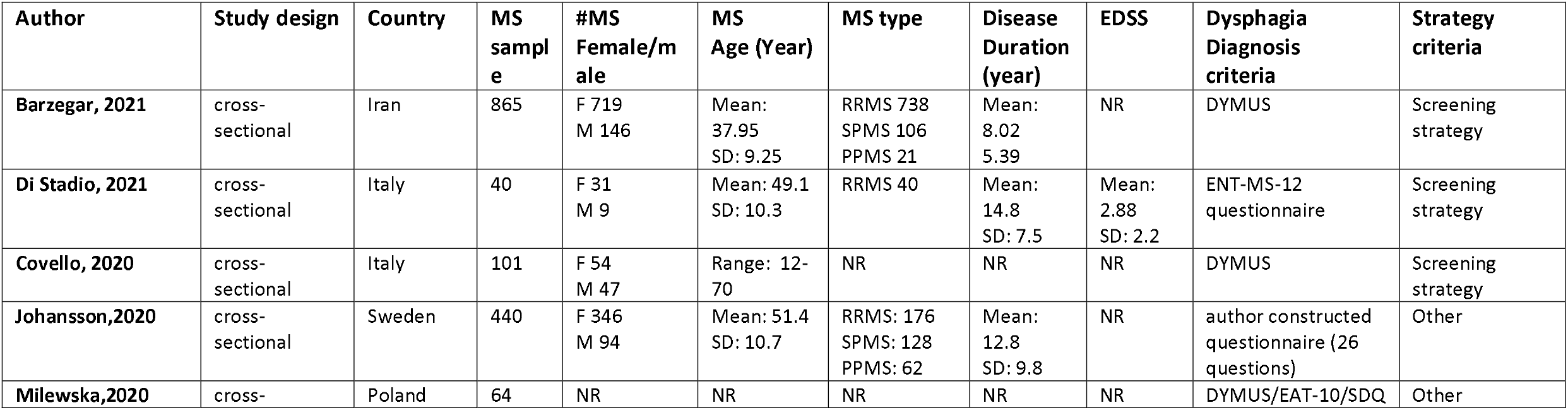
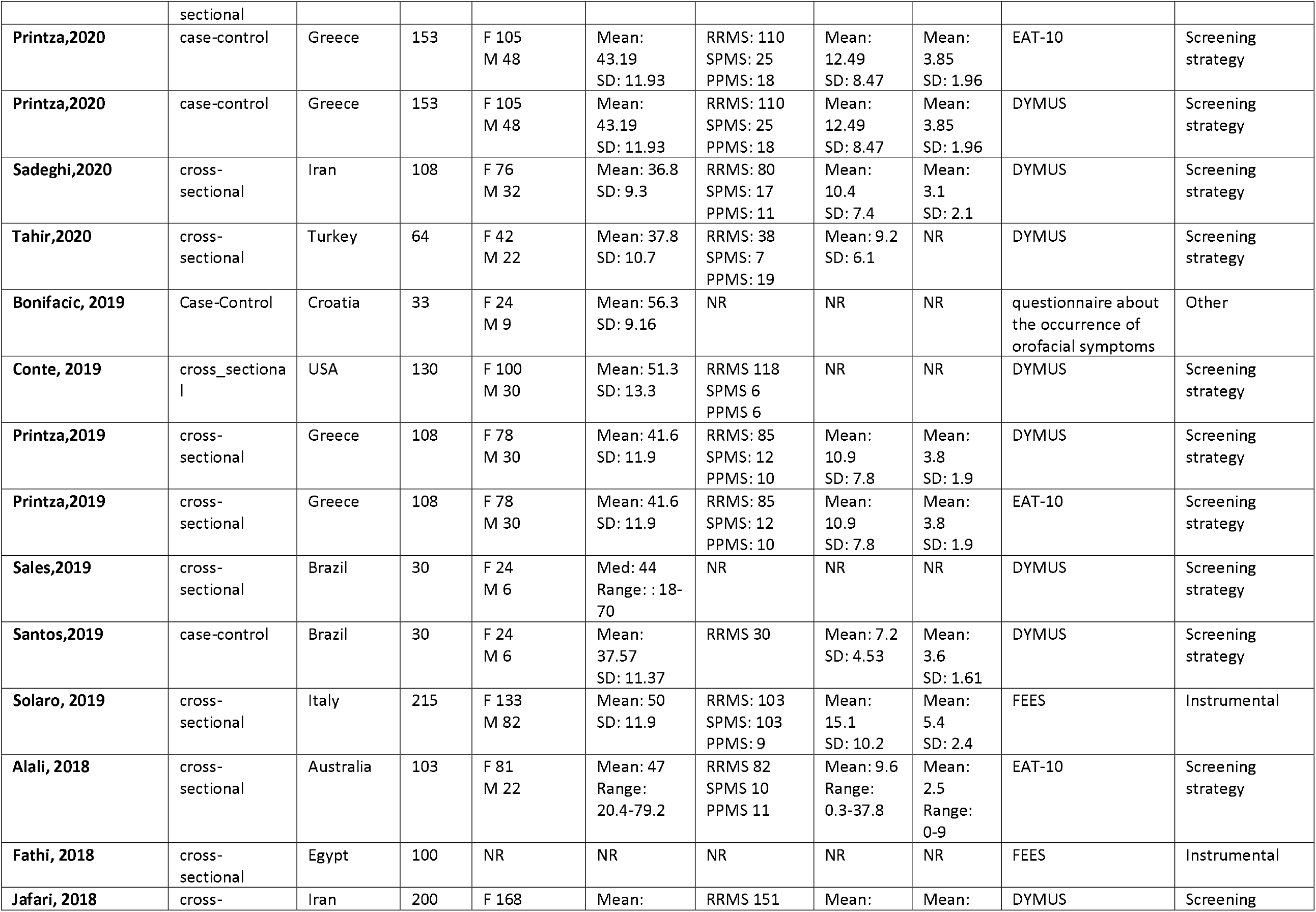
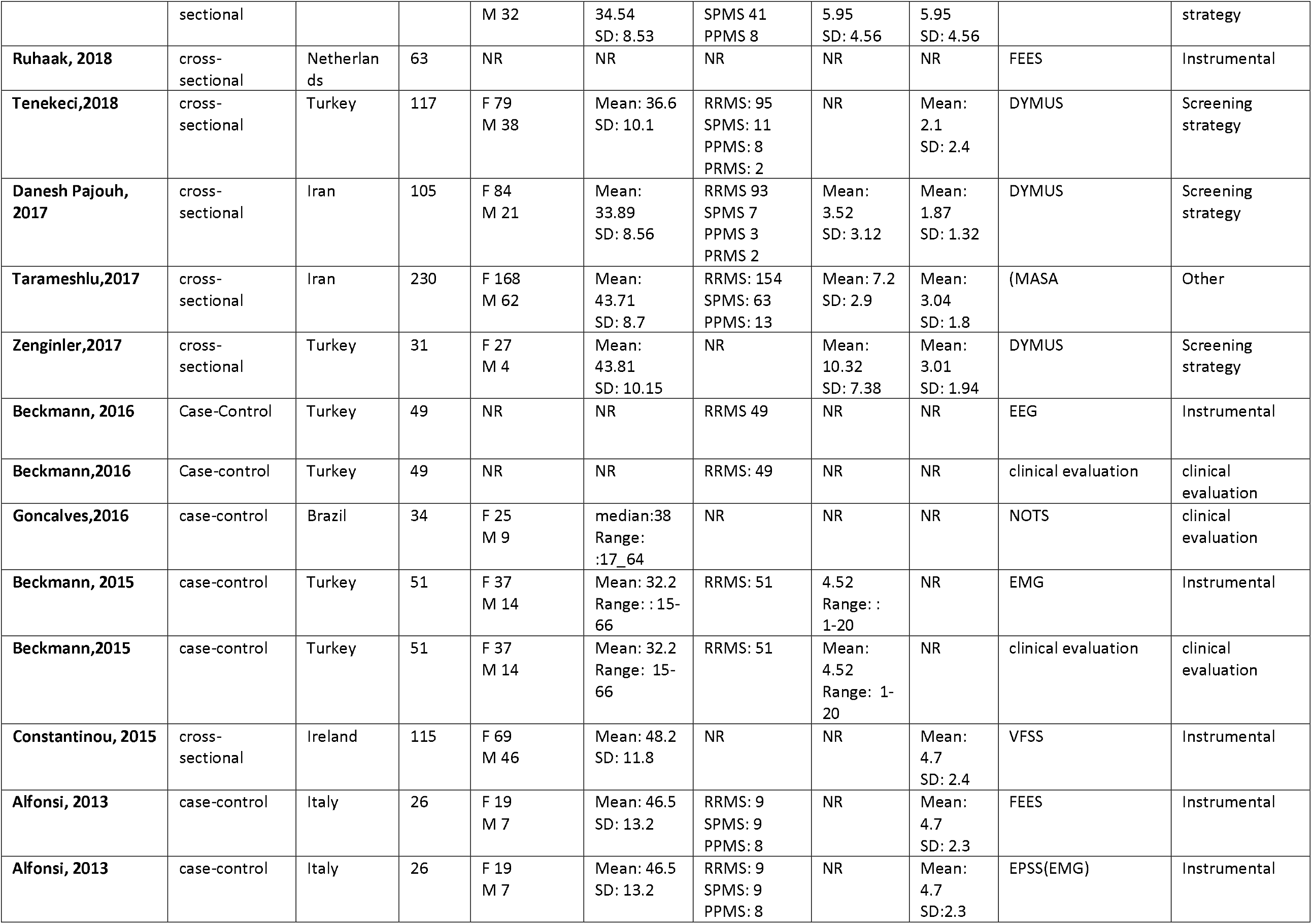
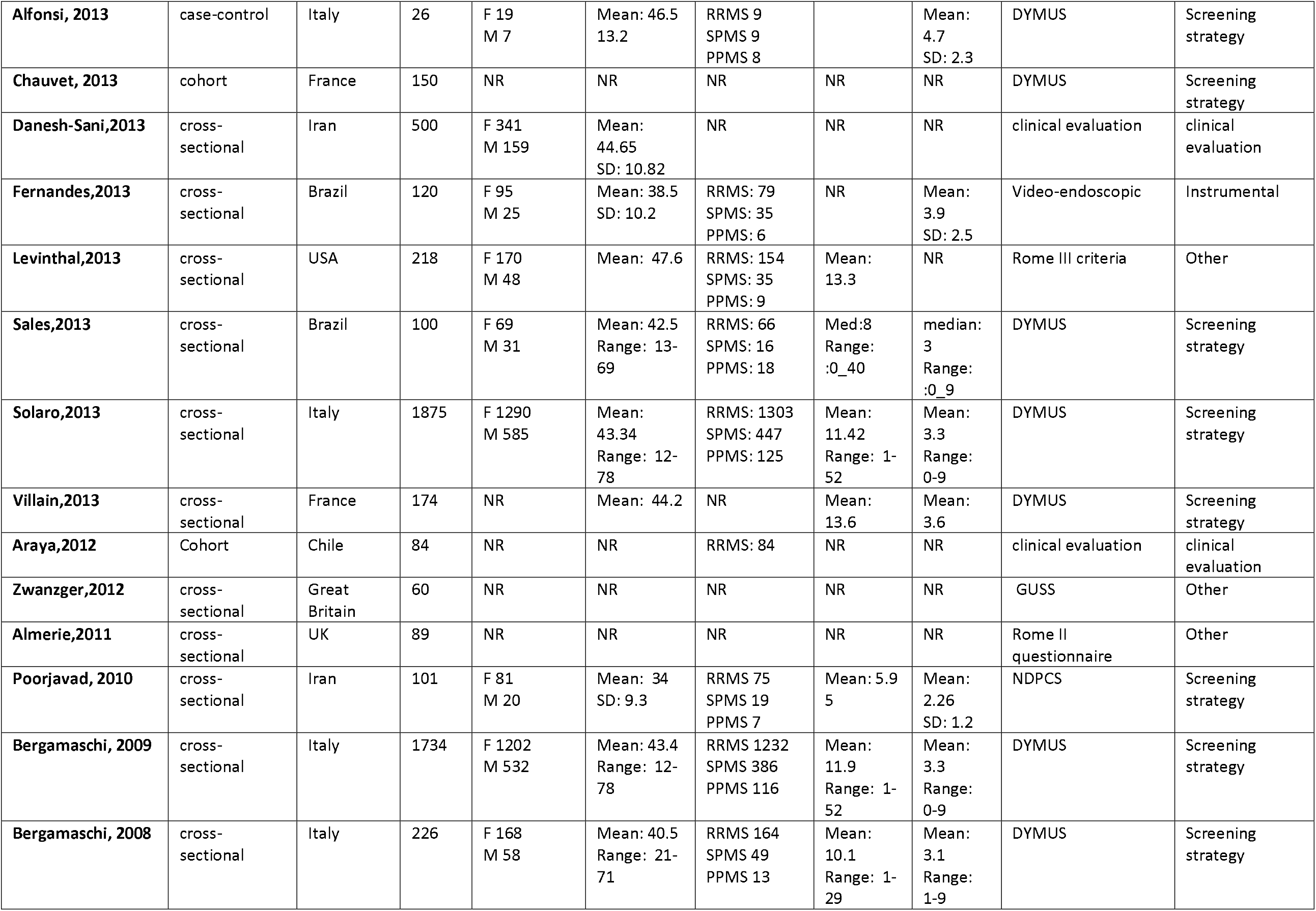
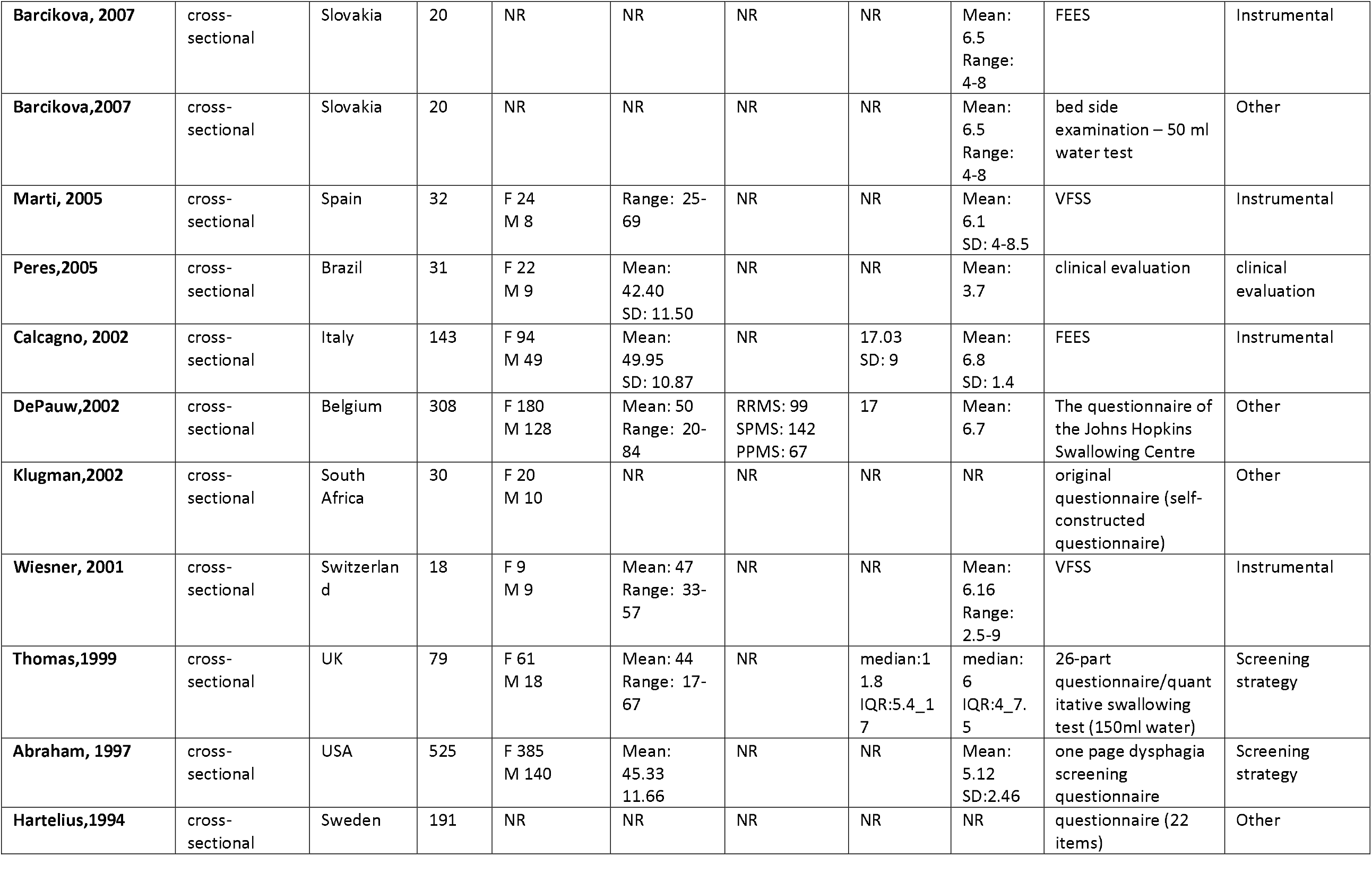
Basic characteristics of included studies

## Discussion

There are a number of studies discussing the prevalence of dysphagia in MS patients. However, all those articles have a number of limitations. For instance, Aghaz et. al. in their article entitled “Prevalence of dysphagia in multiple sclerosis and its related factors: Systematic review and meta-analysis” in 2018, had only reviewed 22 cross-sectional and prospective cohort studies(11) whereas in our study every single article regardless of the study type has been reviewed. Another factor that makes our study stand out is the methodologically rigorous method we had in terms of inclusion criteria. As such, Guan et. al., in a study entitled “Prevalence of dysphagia in multiple sclerosis: A systematic review and meta-analysis” in 2015, claim that around 80% of their analyzed articles had provided an estimate based on self-report screening tests which led to a high level of heterogeneity,(12) while in our study articles with objective and valid methods were included. Our results suggest that 45.3% of MS patients suffer from dysphagia, far higher than that in the general population which is 16-23%. (13)

There are numerous diagnostic methods for dysphagia.(14) In our study we focused on four major methods including instrumental strategies, screening strategies, clinical examinations, and non-instrumental methods. Instrumental methods include video fluoroscopic swallowing examination strategy and fiberoptic endoscopic examination of swallowing strategy. Screening strategies consist of valid and reliable questionnaires designed to screen specifically MS patients for dysphagia, examples of such questionnaires include but are not limited to the Dysphagia in Multiple Sclerosis (DYMUS) questionnaire, the Eating Assessment Tool (EAT-10) questionnaire, the 3-Ounce (90-cc) Water Swallow test, the Dysphagia Screening Questionnaire for MS (DSQMS), and the Northwestern Dysphagia Patient Check Sheet (NDPCS) questionnaire.(8,14–17) Non-instrumental methods are quite uncommon, therefore we coined the term “other methods” to represent studies whose main screening tests were either non-instrumental strategies, or any method other than instrumental, clinical, and screening methods.

Aside from the method that was used to diagnose dysphagia, another important factor in classifying this disease is based on the anatomical location it affects the most. In this manner, dysphagia can be categorized as oropharyngeal, oral, pharyngeal, and esophageal subtypes. However, as one can assume this classification can loosely encompass all the different manifestations of dysphagia.(18–22) Hence, neurologic dysphagia is diagnosed in a patient when the underlying error that leads to dysphagia is within the neural circuits that orchestrate dysphagia rather than related anatomical structures and functional dysphagia is detected in the absence of any anatomical and neuronal abnormalities that might contribute to difficulty swallowing. (23,24) Another aspect of dysphagia is whether or not the patient experiences different levels of difficulty when swallowing different forms of foods such as liquid or solid.(23,25) The aforementioned subtyping is not common although it provides us with perspective into the vast field and various manifestations of dysphagia.

Literature on the association between MS and dysphagia is rapidly growing and previous studies have mentioned dysphagia as a common symptom of MS based on a number of techniques such as screening strategies, instrumental, non-instrumental, and clinical examinations.(5,16,24,26–29) Many articles also categorize dysphagia based on its severity into 5 classes including slight, alarming, mild, moderate, and severe, whereas others may choose to classify this disorder based on the anatomical location that is primarily affected, such as oral, oropharyngeal, pharyngeal, esophageal, etc. there are also neurogenic and functional types of dysphagia that are due to functional disturbances in the swallowing mechanisms rather than its anatomical structures.(18,23) Dysphagia is also reported to have associations with patients’ age, MS type, and disease course. (5)

Dysphagia seems to be strongly connected with the severity of brainstem impairment. Present data reflects the importance of the brainstem in the control of swallowing. The more damage the brainstem sustains, the higher are the chances of developing dysphagia. The aforementioned hypothesis is supported by the role of this critical CNS area in the control of the deglutition mechanism and its neural tract pathways.(25,30) Although the ventromedial reticular formation and the solitary tract nucleus do appear to have a central role in the control of deglutition and its coordination with respiration, the precise location of all the integral parts of the central swallowing pathway has not yet been identified.(3)

Ultimately, we need to focus on the management of dysphagia among MS patients in an effort to prevent its catastrophic side-effects. Two of the of the most common treatment methods include electrical stimulation and the use of botulinum toxin, both of which have shown promise for reducing the swallowing impairment among MS patients. However, there is still argument about whether or not such treatments possess clinical applicability and long-term treatment efficacy and more research is required to enlighten the ambiguity surrounding dysphagia management. (31,32)

The present study has some strengths. First, it is the first methodologically rigorous systematic review and meta-analysis evaluating the prevalence of dysphagia among MS patients with a low risk of publication bias. Second, we managed to categorize the analysis based on different dysphagia diagnostic strategies which has never been done before. However, we had some limitations, too. For example, classifying dysphagia based on its type was not feasible as data was not available in the included articles. Second, we were unable to identify the type of medication that MS patients with dysphagia were receiving. This is an area that might need further research since it can play a major role in identifying those at risk and help apply preventive measures. Third, based on analyzed articles, compensatory strategies utilized by MS patients such as postural changes, and modification of the amount and speed of food presentation were not identified. Identification of such strategies requires close observation of the feeding abilities of patients and can be helpful for better evaluating MS patients’ dysphagia course.

## Conclusion

The results of this systematic review and meta-analysis show that the prevalence of dysphagia in MS patients is significantly higher than the general population. It also provides us with insight into the importance of routine and systemic checkups in MS patients in an effort to prevent the progression of severe comorbidities, especially because dysphagia can lead to aspiration and pneumonia which are both life-threatening in MS patients.

## Data Availability

All of the data will be available for secondary analysis in necessary cases from the corresponding author through email address.

## Abbreviations

EDSS: Expanded Disability Status Scale
MS: Multiple Sclerosis
RRMS: Relapsing-Remitting Multiple Sclerosis
SPMS: Secondary-Progressive Multiple Sclerosis
PPMS: Primary-Progressive Multiple Sclerosis
PRMS: Progressive-Relapsing Multiple Sclerosis
SD: Standard Deviation
IQR: Inter-Quartile Range
Med: Median
F: Female
M: Male
UK: United Kingdom
USA: United States of America
VFSS: Video-Fluoroscopic Swallow Study
EEG: Electroencephalogram
FEES: Fiberoptic Endoscopic Evaluation of Swallowing
EPSS: Electro-Physiological Study of Swallowing
EMG: Electromyography
DYMUS: Dysphagia in Multiple Sclerosis
GUSS: “Gugging Swallowing Screen”
NDPCS: Northwestern Dysphagia Patient Check Sheet
NOTS: Nordic Orofacial Test-Screening
EAT-10: Eating Assessment Tool
ENT-MS-12: Ear, Nose, Throat Multiple sclerosis
MASA: Mann Assessment of Swallowing Ability
Other methods: non-instrumental methods and any method except for instrumental, clinical, and screening strategies

## Conflict of interests

None to be declared

## Funding

None

## Notes

**Conflict of interest:** none to be declared

### Competing Interest Statement

The authors have declared no competing interest.

### Funding Statement

No funding was received.

### Author Declarations

This is a systematic review and the authors declare that all the ethical aspects have been taken care of.

